# Long-term adherence, safety and effectiveness of nusinersen in spinal muscular atrophy patients: a population-based study

**DOI:** 10.64898/2026.03.11.26348135

**Authors:** Karolina Aragon-Gawinska, Nancy-Carolina Ñungo-Garzón, Nuria Muelas, Rafael Sivera, Teresa Sevilla, David Hervás, Inmaculada Pitarch Castellano, Juan F Vázquez-Costa

## Abstract

Nusinersen was the first disease modifying treatment approved for 5q spinal muscular atrophy (SMA). Long-term results of broad populations, particularly for adolescents and adults, remain limited.

We conducted a population-based, ambispective observational study of all SMA patients living in the Valencian Community (Spain) between September 2017 and December 2022 and follow-up until December 2025. Demographic, clinical and motor outcomes using revised SMA Functional Composite Score (SMA-FCR) were collected. Patients were classified as responders or non-responders. The risk for nusinersen discontinuation was assessed with a Bayesian model, and SMA-FCR trajectories with mixed linear regression.

Of 72 patients included, 18 were <12 years old (all treated with nusinersen) and 54 were ≥12 years (28 treated; 26 untreated) at the baseline visit. After a median of 7 years, all patients <12 years were classified as responders versus 68% of patients ≥12 years. Discontinuation rates were 11% in children compared to75% in the older cohort. In patients ≥12 years, reasons for discontinuation included: treatment burden (71%), and loss(53%) or lack of benefit (43 %). Lower baseline SMA-FCR (expEstimate= 0.84 [0.718,0.93], prob:1) and older age (expEstimate=1.028 [1.011,1.055], prob:1) independently predicted higher discontinuation risk. Sustained nusinersen treatment was independently associated with SMA-FCR increase, while untreated and discontinued patients showed slight deterioration over time.

In this long-term population-based study, nusinersen use and persistence was high in children but declined significantly after age 12 due to treatment burden and limited efficacy. However, a proportion of adolescents and adults (those younger and with higher baseline function) experienced sustained benefit.

## Introduction

Spinal muscular atrophy (SMA) is an inherited autosomal recessive disorder caused by biallelic mutations in *SMN1* gene [1], leading to motoneurons loss, which ultimately results in a progressive weakness and atrophy of skeletal muscles. The paralogous *SMN2* gene is one of the strongest disease modifiers, as the number of *SMN2* copies correlates directly with disease phenotype [2]. Classically, four SMA types have been distinguished: SMA type 1 manifesting in the first 6 months of life with rapid motor decline, respiratory insufficiency and high infant mortality in absence of treatment [3]; SMA type 2, appearing between 6 and 18 months of life with patients never achieving independent walking and presenting severe motor disability and scoliosis, ; SMA type 3, where symptoms occur after patients have acquired the ability to walk; and SMA type 4, where symptoms start in the third decade of life [4]. Patients with 2 *SMN2* copies typically develop severe type 1, patients with 3 *SMN2* copies mostly present with SMA type 2 and patients with 4 or more *SMN2* copies usually present as SMA type 3, or less likely, type 4 [2]. With the appearance of disease-modifying therapies, classical SMA types no longer reflect the variety of phenotypes and prognosis, for which reason another functional classification of patients as walkers, sitters and non-sitters is preferred [5].

The approval of nusinersen, an antisense oligonucleotide (ASO) and the first regulatory-approved drug for SMA, has greatly impacted patients’ prognosis and changed the SMA landscape forever. Clinical trials’ data and their open label extensions *(NCT 02594124)* showed a remarkable benefit in survival, motor milestones and motor scales for infants and children up to 12 years old treated with nusinersen [6–10], with only mild adverse events. Subgroup analysis in those studies showed greater benefit in patients receiving treatment earlier. Consequently, a study conducted in presymptomatic patients [11] showed even greater benefit with patients with 2 copies acquiring motor milestones never observed in natural history and patients with 3 copies having a nearly normal motor development [11–13]. Despite this evidence, no randomised trials of nusinersen in older populations have been conducted to date.

Real-world studies brought results to fill the gap on nusinersen’s safety, tolerability and efficacy in adolescents, adults and more severely impaired patients. Thus, a recent meta-analysis showed modest improvements in motor scales (HFMSE, RULM and 6MWT) within these populations [14]. However, moderate-to-high heterogeneity across studies, suggests variability in clinical practice, patient selection, and evaluators’ training. Furthermore, most studies had a median follow-up of less than three years. Long-term data in adolescent and adults remain scarce and conflicting: while some studies suggest sustained improvement or stabilization at 38th months [15], others have observed functional deterioration despite nusinersen treatment [16,17]. Notably, previous research has lacked population-based design and rarely considered treatment withdrawal [15,18] or included untreated patients as an external comparator [19].

Therefore, this study aims to assess the long-term results of nusinersen treatment specifically addressing the existing data gap in adolescent and adult patients through a population-based approach.

## Methods

### Study design

We conducted a single-centre, observational, ambispective, population-based study. The study was performed and reported in accordance with the Strengthening the Reporting of Observational Studies in Epidemiology (STROBE) guidelines for cohort studies [20].

### Population

Currently, all SMA patients living in the Valencian Community (ca. 5 million inhabitants) are being treated and followed up at Hospital la Fe. For this study, we retrospectively reviewed all genetically confirmed SMA patients who attended Hospital La Fe, and included all patients living in the Valencian Community between September 2017 (start of the registry) and December 2022. Prevalent cases as of December 2022 were used to calculate point prevalence. Patients were followed up until December 2025 or until death, inclusion in a clinical trial, switch to another treatment (whichever occurred first). They were excluded from analysis if they lacked either a pre-treatment evaluation in our centre before January 2023; or at least one post-treatment evaluation before the end of follow-up. Demographic, clinical and genetic data of patients were prospectively collected within CUIDAME National Registry [21] following informed consent (see below).

### Procedures

In Spain, nusinersen has been available since March 2018, with access granted in singular cases under Expanded Access Program (EAP) access from 2017. Risdiplam was first available under compassionate use in 2020 for selected patients with no access to nusinersen (complex spines) and has become widely available under reimbursement since January 2023. Patients were treated according to the Spanish treatment protocol from April 2018 and its posterior revision, based on the Spanish Delphi consensus [22]. This protocol establishes inclusion, exclusion and discontinuation criteria, together with follow-up recommendations (supplementary material). Nusinersen treatment was indicated by one expert neuropediatrician (IPC) in paediatric (<15 years old) and one expert neurologist (JFVC) in adolescent/adult patients, after discussing potential benefits and risks of treatment with the patients (or legal representatives). Nusinersen was delivered as per label by lumbar puncture in 4 loading-dose injections within 2 months and posteriorly every 4 months. Radiological (ultrasound, CT or fluoroscopy) guided lumbar puncture was used in patients with complex spines, including spinal fusion, as previously published [23].

Both treated and untreated patients were evaluated every 4-12 months (usually every 8 months), using motor scales according to their age (supplementary material). Although Hammersmith Functional Motor Scale - Expanded (HFMSE) and Revised Upper Limb Module (RULM) were initially developed for children with SMA type 2 and 3, they were later validated in adolescents and adults [24]. However, these scales exhibit floor effects in non-sitters and low function sitters, as well as ceiling effects in walkers [25]. Ambulatory patients were additionally assessed with the six-minute walk test (6MWT), which has beem validated in both children and adult SMA patients [26]. To minimise the floor and ceiling effects of HFMSE and RULM and to enable comparison across different phenotypes, the Spinal Muscular Atrophy Functional Composite Score Revised (SMA-FCR), merging all three scales, was calculated [17]. Adverse events were recorded at each visit.

After two years of treatment (or before if requested by patients) and at every follow-up visit, the continuation of nusinersen was re-evaluated. Clinical benefit was evaluated using both motor and functional scales, as previously reported [25]. Patients who showed lack of benefit, as assessed by the clinician and perceived by the patient, or those with an unfavourable risk-benefit balance (considering side effects and patient burden) were proposed to discontinue treatment. All therapeutic decisions were made in agreement with patients. The reasons for discontinuation of treatment were recorded in medical records and retrospectively classified into three categories: (1) lack of perceived benefit (never experienced benefit, as evaluated by clinician and perceived by the patient), (2) loss of an initial benefit, and (3) treatment burden (including adverse events related to the treatment or its administration, but also inconveniences of repeated admissions for lumbar punctures). One single patient could have more than one reason for discontinuation (e.g. loss of benefit and treatment burden).

At the end of follow-up, adolescent and adult patients were classified into three groups: sustained responders (those reporting some benefit and showing a sustained improvement, considered as clinically significant), partial responders (those showing an initial improvement in motor scales, but losing it subsequently, or reporting benefit, that was not appreciated in motor scales) and non-responders (those who never reported or showed any consistent improvement in motor scales).

#### Ethics

All patients signed informed consent for participation in CUIDAME registry. The study was approved by the bioethical committee of Hospital la Fe (2019-027-1).

#### Statistics

Data were summarized as means, standard deviations, medians, and first and third quartiles for the continuous variables, and as relative and absolute frequencies for the categorical variables. Exploratory descriptive analyses and graphs were used to assure the quality of the data and describe the progression of SMA-FRC. Trendlines were estimated using smoothing splines. The first visit was the baseline visit right before starting nusinersen in treated patients or the first overall visit in untreated patients. The last visit was the last available visit before censoring (starting clinical trial or starting a new treatment). For the purposes of this study, patients were divided into 2 subgroups according to age at first visit: <12 years (children) and ≥12 years (adolescents and adults). This age was chosen considering the natural history of the disease (which considerably changes in adolescence) and our experience with nusinersen.

In patients ≥12 years a Bayesian model was developed to assess the independent effect of age, sex and baseline SMA-FCR in the risk of nusinersen discontinuation. Moreover, a mixed linear regression model was used to compare SMA-FCR trajectories with time in the three subgroups of patients (untreated, treatment discontinuation and sustained treatment), adjusting for age and sex. All the statistical analyses and graphs were performed with R software (version 4.5.2).

## Results

We identified 107 SMA patients visited at Hospital la Fe. Ninety-eight of them were living in the Valencian region in December 2022, so the estimated point prevalence was 98 cases/5,059,00 inhabitants = 1,94 cases/100.000 inhabitants.

Of the 107 patients, 28 were excluded due to the lack of a baseline visit in our centre before January 2023 and 7 because they lacked a follow-up visit. Consequently, 72 patients were finally included in the study and were followed up for a median of 7 years (IQR 5.5, 7.4). Of those, 18 were <12 years at the onset of treatment, while 54 were ≥12 years (Figure 1).

**Figure 1.**
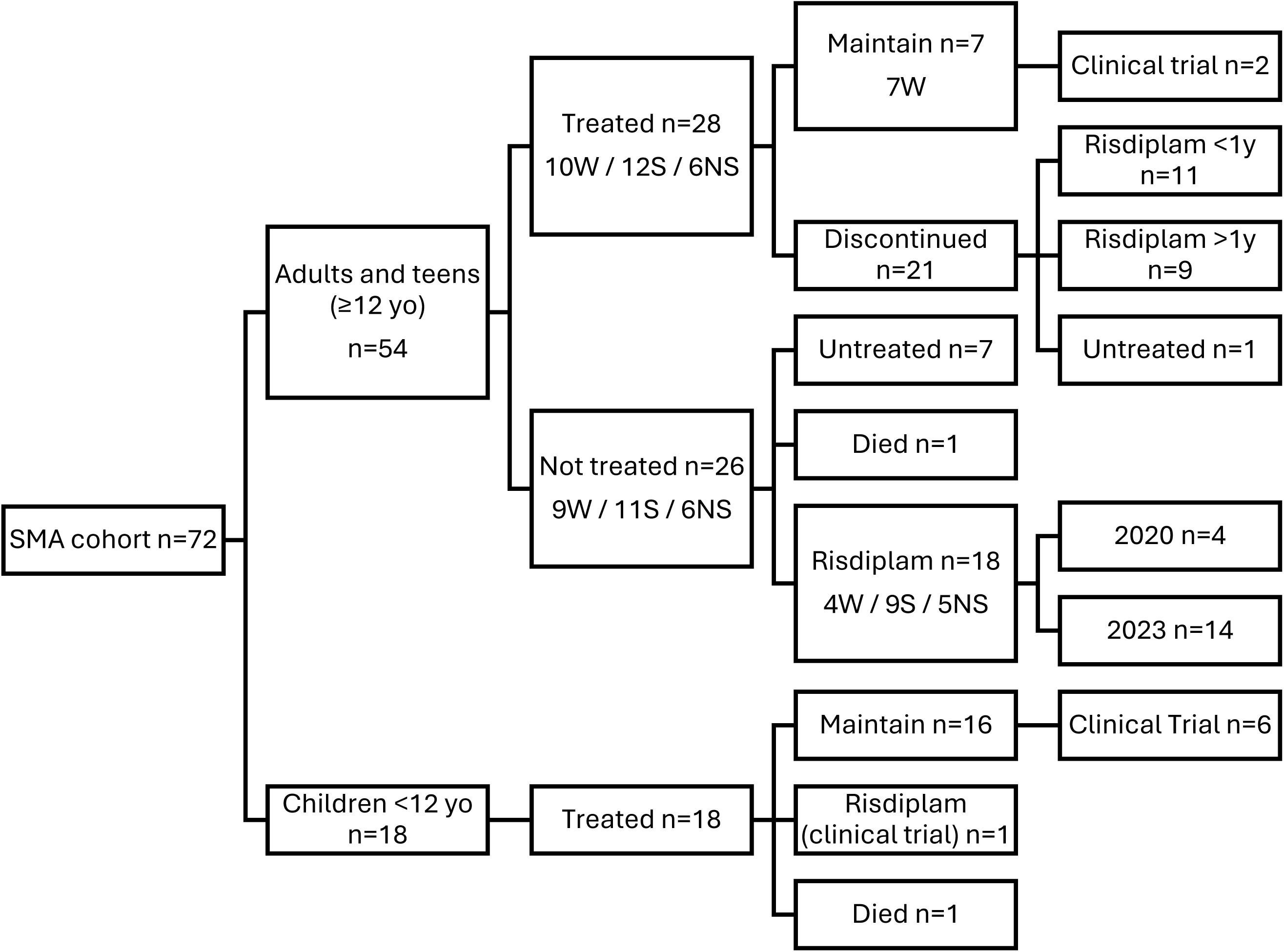
Flow chart of included patients. W – walker, S – sitter, NS – non-sitter; Risdiplam <1y means switch to risdiplam within one year of nusinersen discontinuation; Risdiplam >1y means risdiplam started more than 1 year after nusinersen discontinuation.

By December 2022, 46 SMA patients (64%) had been treated with nusinersen and 26 patients remained untreated. Baseline characteristics of patients can be found in Table 1. Briefly, all children <12 years were treated (22% with radiological guidance) vs only 28 of 54 (54%) patients ≥12 years (46% with radiological guidance). Baseline characteristics of untreated vs treated patients ≥12 years were similar in terms of *SMN2* copies, SMA type and functional status, although untreated patients were older and more frequently female (Table 1). The reasons for not starting nusinersen treatment included: lack of intrathecal access despite radiological guidance in four patients (15 %), not meeting reimbursement criteria in five individuals (19 %) (two lacked residence permit, one had minimal motor function and ≥16 hours of non-invasive ventilation, and twoexhibited normal results in motor scales), and patient’s preference in 17 cases (61%). Of those 26 untreated patients, 18 had started treatment with risdiplam at the end of follow-up, while seven continued untreated and one died due to respiratory insufficiency.

**Table 1.** Baseline characteristics of patients included in the study according to nusinersen treatment.

|  | Nusinersen treated patients |  |  | Untreated patients<br>(>12 yo)* |
| --- | --- | --- | --- | --- |
|  | All treated | <12 yo | ≥12 yo |  |
| n | 46 | 18 | 28 | 26 |
| Age in years median<br>(IQR)** | 14.95 (6.45, 32.08) | 3.02 (1.7, 6.9) | 27.2 (16.33, 45.25) | 35.08 (25.11, 42.85) |
| Gender F/M n (F%) | 26/20 (56%) | 14/4 (78%) | 12/16 (43%) | 17/9 (65%) |
| SMA type, n (%) |  |  |  |  |
| I | 2 (4%) | 2 (11%) | 0 | 0 |
| II | 19 (41%) | 9 (50%) | 10 (36%) | 11 (42%) |
| III | 24 (52%) | 6 (33%) | 18 (64%) | 14 (54%) |
| IV | 0 | 0 | 0 | 1 (4%) |
| Presymptomatic | 1 (2%) | 1 (6%) | 0 | 0 |
| SMN2 copies |  |  |  |  |
| 1 *** | 1 (2.17 %) | 0 (0 %) | 1 (4%) | 0 (0 %) |
| 2 | 3 (6.52 %) | 2 (11.11 %) | 1 (4%) | 0 (0 %) |
| 3 | 31 (67.39 %) | 14 (77.78 %) | 17 (61%) | 17 (65.38 %) |
| 4 | 11 (23.91 %) | 2 (11.11 %) | 9 (32%) | 9 (34.62 %) |
| Functional status |  |  |  |  |
| Non-sitter | 6 (13.04 %) | 0 (0 %) | 6 (21.43 %) | 6 (23.08 %) |
| Sitter | 24 (52.17 %) | 12 (66.67 %) | 12 (42.86 %) | 11 (42.31 %) |
| Walker | 16 (34.78 %) | 6 (33.33 %) | 10 (35.71 %) | 9 (34.62 %) |
| Baseline SMA-FCR | 28.36 (12.22, 53.51),<br>n=38 | 29.98 (23.94, 51.2),<br>n=10 | 28.12 (12.07, 52.37),<br>n=28 | 19.36 (9.23, 77.7),<br>n=25 |
| Scoliosis | 22 (47.83 %) | 6 (33.33 %) | 16 (57%) | 15 (57.69 %) |
| NIV use | 9 (19.57 %) | 2 (11.11 %) | 7 (25%) | 7 (26.92 %) |
| Radiology assisted lumbar puncture n (%) | 17 (37%) | 4 (22%) | 13 (46%) | NA |
| Follow-up (months), median (IQR) | 83.88 (75.22, 89.07) | 83.41 (56.03, 90.89) | 84.26 (80.22, 88.2) | 85.93 (55.6, 89.07) |
| Treatment duration (months), median (IQR) | 59.02 (34.94, 83.71) | 83.41 (56.03, 90.89) | 48.7 (22.57, 75.88) | NA |
| Deceased, n (age in years) | 1 (12) | 1 (12) | 0 | 2 (38, 74) |

\*All untreated patients were older than 12 yo.
\*\* Age at baseline visit
\*\*\*SMN1 point mutation

After a median treatment duration of 83.4 months (IQR 56, 90.9) all children <12 years old were classified as responders at their last visit: three as partial and 15 as sustained responders. Two children discontinued nusinersen treatment during follow-up (Figure 1): one patient died at the age of 12, after 22 months of therapy due to upper gastrointestinal bleeding, probably unrelated to treatment or SMA; another, discontinued nusinersen to participate in a clinical trial with risdiplam. Additionally, six children treated with nusinersen were included in clinical trials and their follow-up SMA-FCR data was censored. Despite this limitation, the SMA-FCR in patients <12 years showed an improvement during the first two years of treatment followed by stabilization (Figure 2).

**Figure 2.**
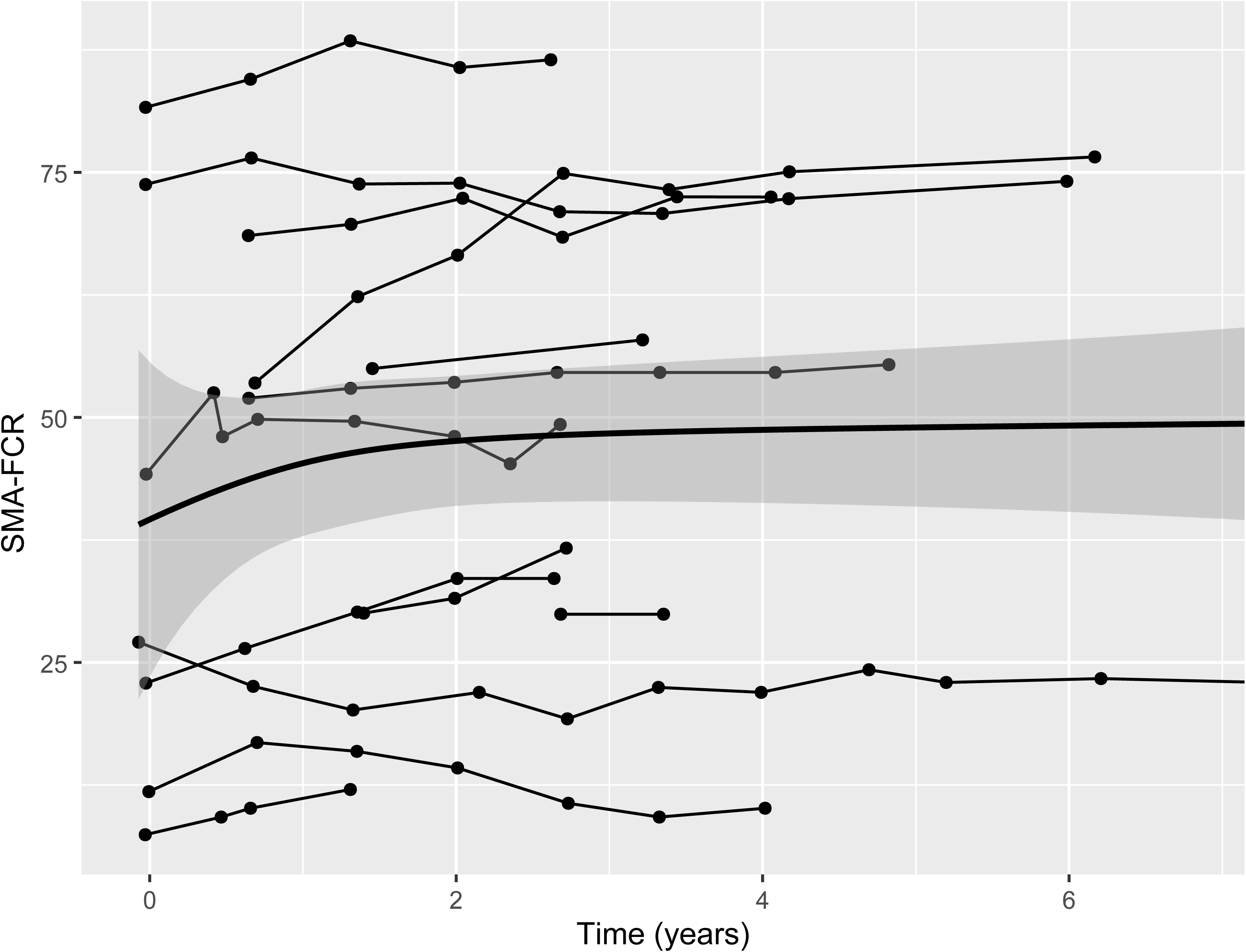
Individual trajectories of SMA-FRC scores in SMA patients: <12 years old treated with nusinersen; SMA-FCR: Spinal Muscular Atrophy Functional Composite Score Revised.

In contrast, after a median treatment duration of 48.7 months (IQR 22.6, 75.9) in patients ≥12 years old, nine patients (32%) were classified as non-responders, 12 (43%) as partial responders and seven (25%) as sustained responders. Consequently, only seven patients (25%) remained treated with nusinersen (one of them with radiological guidance, none of them NIV users) at the end of follow-up (Table 2). The remaining 21 patients stopped the treatment after a median of 35 months (IQR 22.4, 52.7). Reasons for discontinuation included: treatment burden reported by 15 patients (71%), loss of benefit in 11 (53%) and/or lack of benefit in nine patients (43 %). After discontinuation, 11 patients switched directly to risdiplam and nine discontinued treatment and were treated with risdiplam more than one year after discontinuation. One patient stopped treatment and continues untreated until now.

**Table 2.** Demographic and clinical characteristics of adolescent and adult patients treated with nusinersen. SMA-FCR: Spinal Muscular Atrophy Functional Composite Score Revised.

|  | Nusinersen discontinued<br>(>12 yo) | Nusinersen maintained (>12<br>yo) |
| --- | --- | --- |
| n | 21 | 7 |
| Age in years median (IQR)* | 29.54 (15.24, 50.17) | 22.35 (19.48, 29.58) |
| Gender F/M n (F%) | 11/10 (52%) | 1/6 (15%) |
| Follow-up in months, median (IQR) | 86.37 (80.9, 87.97) | 84.23 (79.6, 87.41) |
| Nusinersen treated, n (%) | 21 (100%) | 7 (100%) |
| Nusinersen treatment duration (months)<br>median (IQR) | 34.53 (22.37, 52.73) | 84.23 (79.6, 87.41) |
| SMN2 copies |  |  |
| 1** | 1 (4.76 %) | 0 (0 %) |
| 2 | 1 (4.76 %) | 0 (0 %) |
| 3 | 14 (66.67 %) | 3 (42.86 %) |
| 4 | 5 (23.81 %) | 4 (57.14 %) |
| Functional status |  |  |
| Non-sitter | 6 (28.57 %) | 0 (0 %) |
| Sitter | 12 (57.14 %) | 0 (0 %) |
| Walker | 3 (14.29 %) | 7 (100 %) |
| Baseline SMA-FCR score | 19.53 (9.51, 33.42), n=21 | 69.58 (69.14, 71.42), n=7 |
| Scoliosis | 13 (61.9 %) | 3 (42.86 %) |
| NIV use | 7 (33.33 %) | 0 (0%) |
| Radiological guided lumbar puncture | 12 (57.14 %) | 1 (14.29 %) |
| Response to nusinersen (n) | 7 | 21 |
| No | 0 (0 %) | 9 (42.86 %) |
| Partial | 2 (28.57 %) | 10 (47.62 %) |
| Responder | 5 (71.43 %) | 2 (9.52 %) |

\* Age at nusinersen's treatment onset or first visit
\*\*SMN1 point mutation

Compared with patients maintaining nusinersen treatment, those who discontinued therapy were older, more frequently female (52% vs. 15%), had lower SMA-FCR and required more frequently radiological lumbar puncture (Table 2). Moreover, all patients discontinuing nusinersen had started treatment either after the age of 40 years old or with a baseline SMA-FCR lower than 50 (Supplementary Figure 1). This association was confirmed in the multivariable Bayesian model, where older age (expEstimate= 1.028 [1.011, 1.055], prob: 1) independently increased the risk of discontinuation, while male sex (expEstimate= 0.148 [0.005, 2.41], prob: 0.902) and higher SMA-FRC scores (expEstimate= 0.84 [0.718, 0.93], prob: 1) reduced that risk (Supplementary Table 1).

In patients ≥12 years maintaining nusinersen treatment at last follow-up (n=7), a mild increase in SMA-FCR during the first 2 years of treatment followed by stabilization was observed (Figure 3a), while those who discontinued treatment (n=21) exhibited stability in SMA-FCR scores (Figure 3a). Similarly, SMA-FCR remained stable in 26 untreated patients (Figure 3b). The mixed lineal model (Figure 4) confirmed that patients maintining nusinersen treatment showed a sustained improvement in SMA-FCR scores (+0.97 points per year), while patients discontinuing nusinersen showed similar trajectory to those never treated.

**Figure 3.**
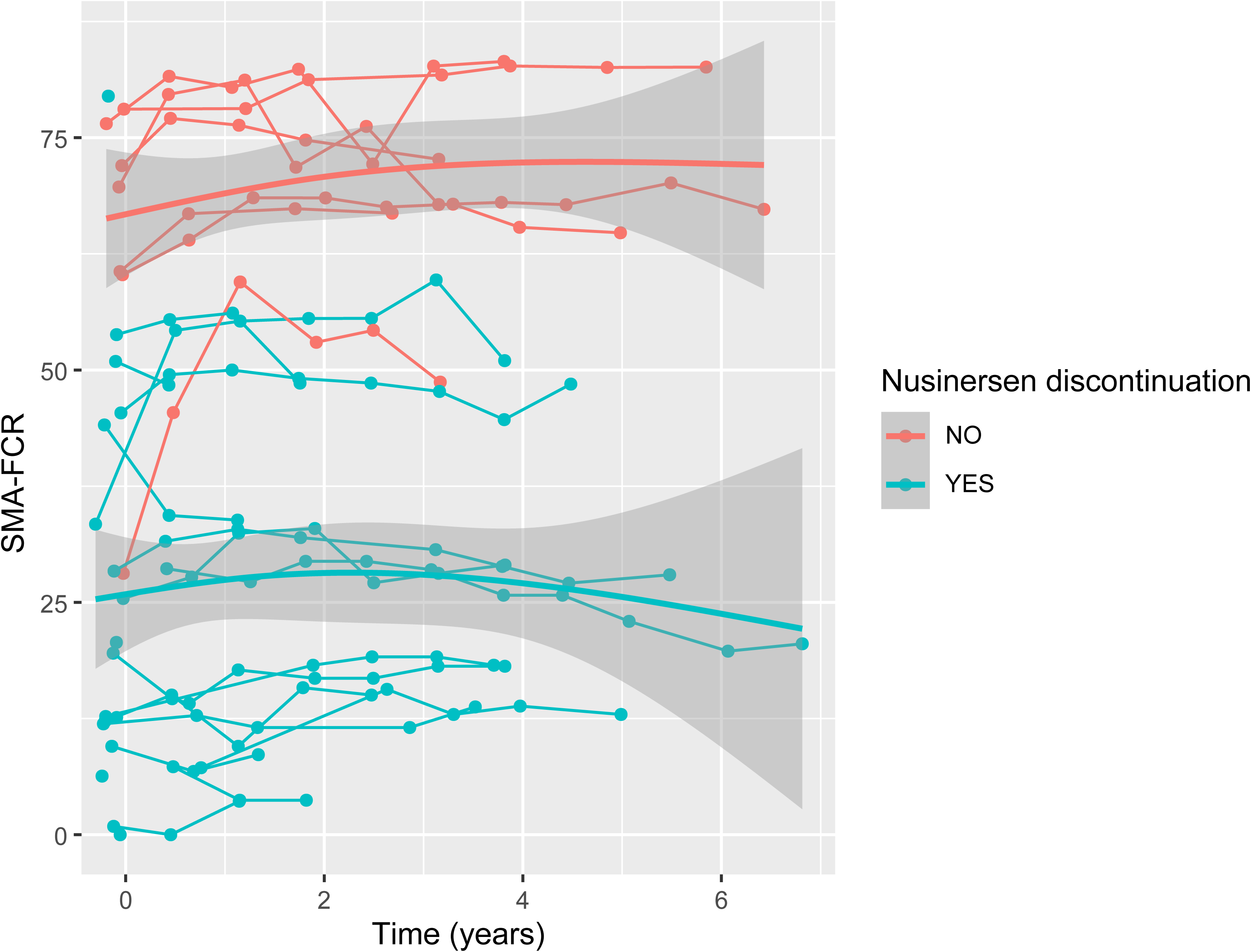
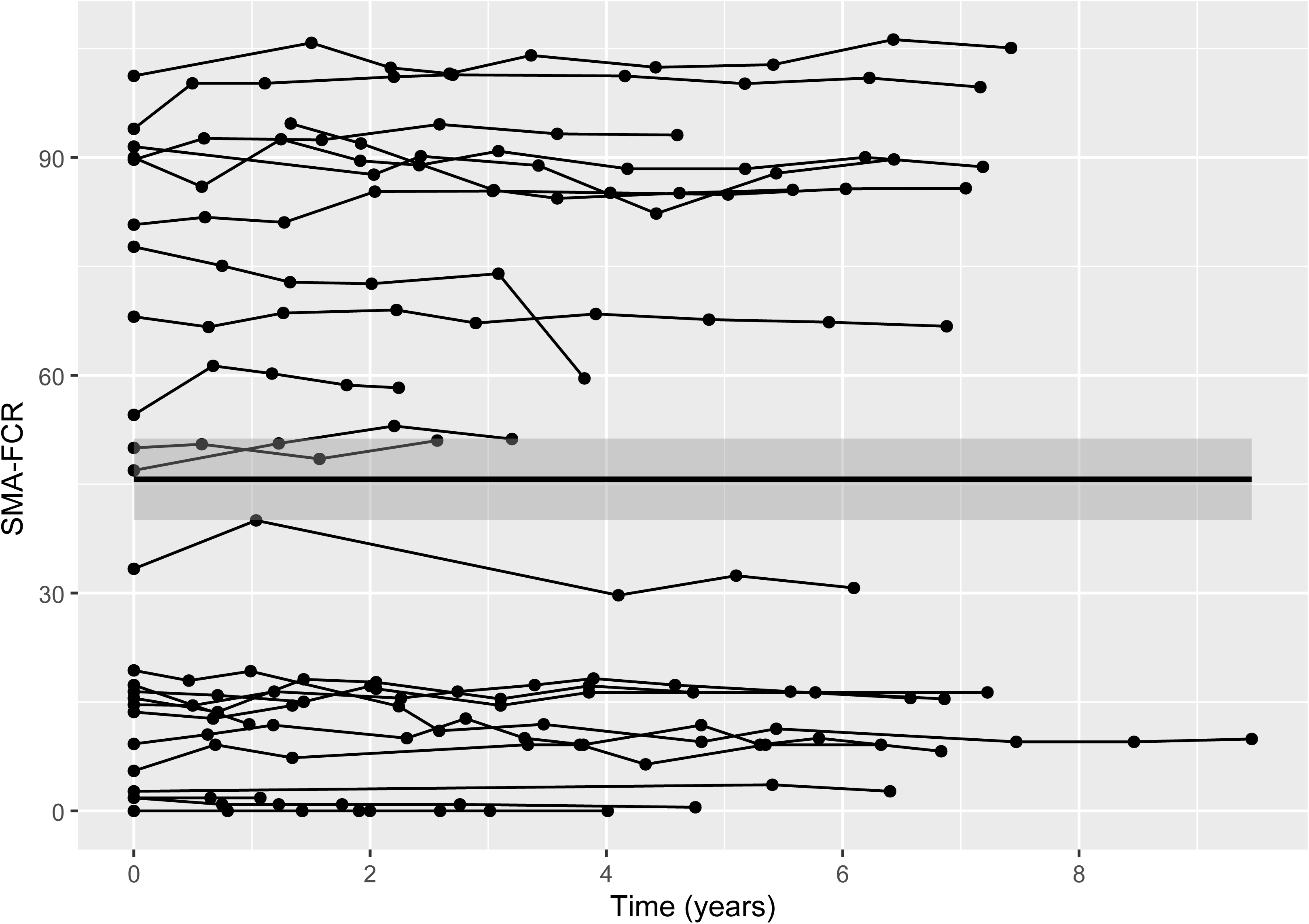
Individual trajectories of SMA-FRC scores in SMA patients ≥12 years old: (a) treated with nusinersen according to its maintenance or withdrawal (b) untreated; SMA-FCR: Spinal Muscular Atrophy Functional Composite Score Revised.

**Figure 4.**
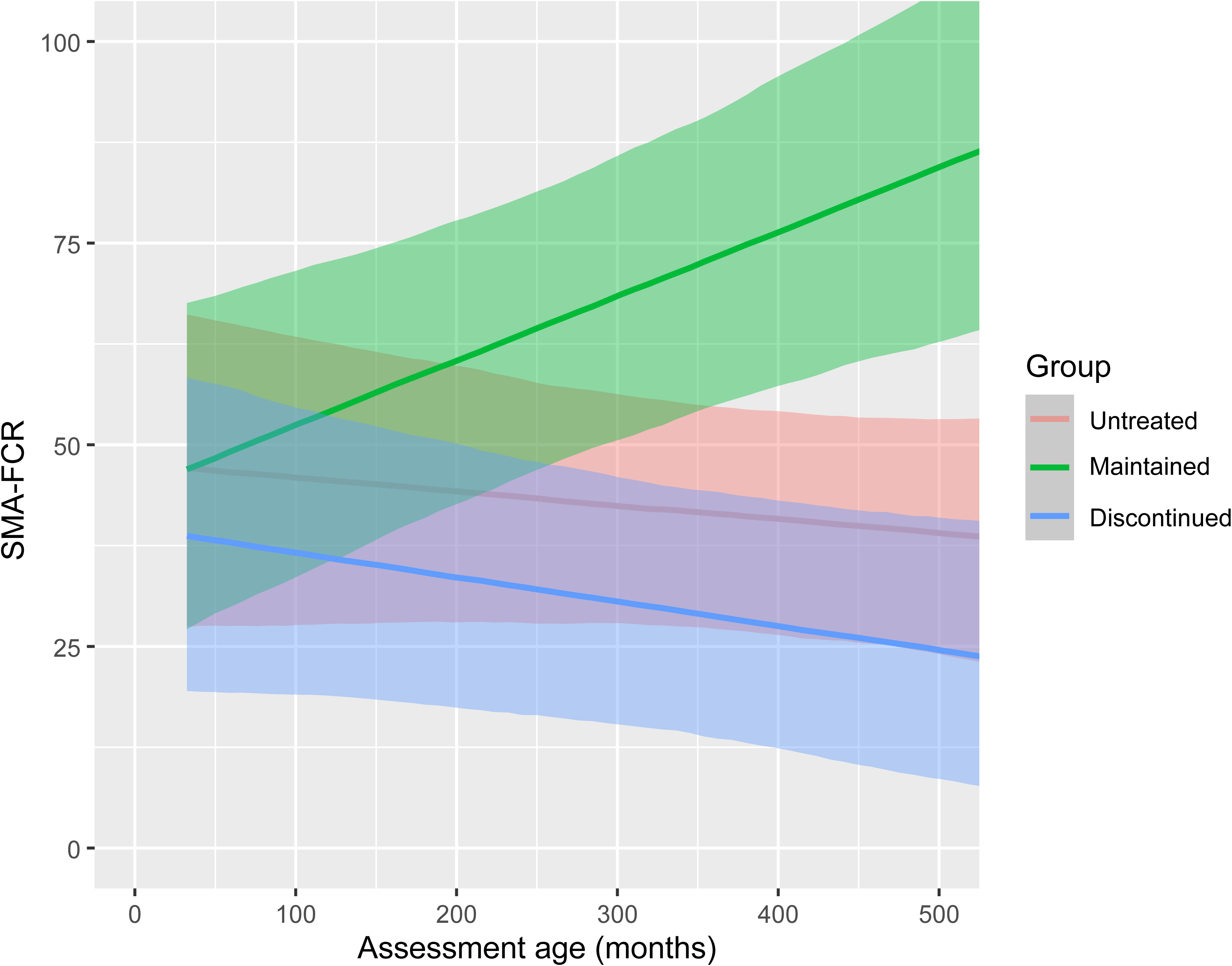
Trajectories of SMA-FRC scores in untreated SMA patients ≥12 years compared to those with sustained nusinersen treatment and those who discontinued nusinersen, based on the mixed linear model. SMA-FCR: Spinal Muscular Atrophy Functional Composite Score Revised.

All patients ≥12 years old treated with nusinersen presented mild AEs, primarly related to the administration procedure. Additionally, two adult patients developed a moderate AE (neurogenic bladder) during the treatment period, which was attributed to nusinersen. In one of them, symptoms partially resolved following discontinuation of the treatment.

## Discussion

This population-based study provides a comprehensive, longitudinal evaluation of nusinersen use, safety and effectiveness in a real-world setting. We demonstrate that while overall long-term nusinersen achieves sustained motor improvements in SMA patients, persistence and risk-benefit ratios vary significantly by age and baseline functional status. Within our region, we estimated a prevalence of 1.94 cases per 100.000 inhabitants, which aligns closely with recent estimates from Italy [27] and remains within the establishedprevalence range for European countries – typically 1-2 cases per 100.000 inhabitants [28]. This suggests that our cohort is highly representative of the broader SMA population, enabling extrapolation of the results to other regions, particularly in Spain.

Population-based studies provide more-balanced results than centre-based studies, that restrict inclusion to treated patients and are particularly susceptible to selection bias and confounding by indication [29]. Patients who receive treatment in referral centres often differ systematically from untreated patients in terms of disease severity, comorbidities, or motivation, which can lead to overestimation of treatment effectiveness [30]. This is especially problematic in observational studies, where treatment assignment is non-random and influenced by clinical judgment, patient preferences, or centre-specific standards of care practices.

Up to now, population-based studies to assess the effectiveness of nusinersen have been reported only in children. In Sweden, nusinersen was found to increase overall survival in SMA type 1 children [31]. In the Netherlands, an improvement of motor function in 72% and stabilization in another 18% of the symptomatic children treated under 9.5 years old was found [32], which differed from the natural disease course found in a matched retrospective cohort. Our results in children are very similar to that study in terms of use (100% in our study versus 89% in the Dutch study), safety (good tolerance) and effectiveness, with > 90% of responders found in both studies and motor function improving up to two years after treatment onset, followed by stability. This resulted in high persistence of treatment in both studies with few discontinuations, and none of them due to adverse events. The favourable motor outcomes observed in these cohorts also align with the results of open label and real-world centre-based studies [17,33–37].

However, our findings reveal less favourable outcomes in patients starting nusinersen after 12 years of age. Firstly, only 54% of patients aged ≥12 years started treatment with nusinersen between 2018 and 2022, and patient’s preference was the main reason to not initiate treatment. In our experience, the uncertainty about safety and effectiveness in the adulthood made many patients reluctant to receive an intrathecal treatment and many preferred to wait for an oral option. This is in line with studies reporting the preferences of SMA adults for oral treatments compared to intrathecal or intravenous ones [38].

Secondly, 32% of adolescents and adults were classified as non-responders with 75% discontinuing nusinersen after a median of 34 months. While these figures align with previous US claims analyses [39,40], they contrast with centre-based studies, where discontinuation is infrequently reported or ranges from 0-16% with only one study reporting 33% rate [14]. Such discrepancies raise questions about the influence of possible publicationor referral bias in centre-based studies, shorter follow-up, and differences in the availability of alternative treatments. For example, another centre-based study reported 57.7% patients being switched to risdiplam [41].

Notably, 71% of patients in our study reported treatment burden as a primary driver for discontinuation. In many patients, particularly with coexisting severe scoliosis, the invasive nature of repeated intrathecal administrations may outweigh the perceived benefit. This is further reflected by 39% of patients who transitioned to risdiplam once it became available. However, the introduction of an oral alternative was only one of the possible reasons, as 35% of patients discontinued nusinersen prior to having access to risdiplam. Furthermore, reports of a lack of benefit (43%) and a loss of benefit (53%) suggest that long-term efficacy may evolve over time. Interestingly, the mixed model showed no significant differences in SMA-FCR trajectories between patients discontinuing nusinersen and untreated patients, suggesting that the decision to stop treatment was related with lack or loss of clinical benefit.

While some centre-based studies suggested large improvements in adolescents and adults [14,41–44], long-term open-label clinical trial [45](SHINE study) and other large observational study [16] have shown that, after 3 years of follow-up, adolescent and adult patients start worsening in motor scales despite nusinersen treatment. It is often argued that, in the context of a neurodegenerative condition, long-term stability or deceleration of declineis a therapeutic response. While we overall agree with this statement, our study shows that extended periods of stability in motor scales also ocurredwithin our untreated patients group. This observation suggests that long *stability* periods may be part of the natural history of the disease in adult patients.

On the other hand, we observed an improvement in SMA-FCR scores followed by a sustained stability in 25% of adolescents and adults, which is unlikely in the natural history of the disease. The mixed model analysis further confirmed a different SMA-FCR trajectory compared to untreated patients. This suggests that nusinersen remains a viable option for a subset of adolescent and adult patients; identifying these individuals is essential to optimizing the risk-benefit ratio of the treatment.

In our cohort, adolescents and adults who maintained long-term treatment were predominantly male, younger than 40 years, ambulatory, and presented with mild scoliosis, and a robust initial response. Furthermore, the multivariable model identified older age, lower SMA-FCR scores and female sex as independent predictors of discontinuation. While higher baseline motor function and ambulatory status have been consistently associated with superior outcomes in adults [16,19,46,47], this could suggest a greater functional reserve, reflecting the density and viability of remaining motor units, is a prerequisite for translating nusinersen mechanism of action into clinically meaningful improvements.

While age at treatment initiation is critical for the response in children, its influence in older patients remains debated [16,19,46,47]. Interestingly, sex has not been previously linked to treatment efficacy; the association of age and female sex with nusinersen discontinuation found in our study probably reflects the greater burden of lumbar punctures with aging and the fact that female sex is a well-documented risk factor for post-lumbar puncture headache [48].

In the absence of reliable predictive or response biomarkers in adolescent and adults [49], variables such as age, sex and baseline motor function should be considered for personalising therapeutic decision and managing patient expectations. Our finding reinforce the growing body of evidence that a significant proportion ofSMA patients face unmet needs, even with the availability of disease-modifying therapies [50]. This highlights the need for exploring new therapeutic interventions. Since most patients who discontinued nusinersen pointed to treatment burden as the main factor, considering alternatives like oral treatments, intrathecal port devices (*NCT06555419*) or less frequent, annual intrathecal dosing (*NCT07221669*) could enhance persistence. Moreover, new options should also be explored to reduce the high economic burden of drugs for patients who show little functional gain from therapy.

### Strengths and limitations

Limitations include the single-centre nature of the follow-up, which might introduce specialist preference bias, though the motor scale results suggest this bias was not relevant. Additionally, while the sample size allowed for multivariable analysis, comparisons between subgroups should be interpreted with caution. Finally, the evaluation of treatment efficacy remains a challenge, particularly in adults and low-function patients [5]. The use of SMA-FCR in this study has allowed for a better comparison of broad cohorts, but still only considers motor function, disregarding bulbar, respiratory or fatigue evaluation, and does not reflect patient perspective [25,51]. Thus, SMA-FCR might be insufficient to capture the complexity of possible treatment’s benefits.

A major strength is the population-based design, which limits the potential bias toward more motivated and proactive individuals. By including data on patients who withdrew from or never initiated treatment, we provide a more realistic reflection of nusinersen’s real-world impact than studies that focus only on those who remain on therapy.

## Conclusion

In this long-term, population-based study, nusinersen persistence was high in children but substantially low after the age of 12. While treatment discontinuation was primarly driven by the high procedural burden and limited perceived effectiveness, the subset of patients who maintained nusinersen treatment showed sustained benefit. Among adolescents and adults, younger age and higher baseline function were key predictors of a more favourable outcome. Integrating these factors into clinical decision-making will help to personalise therapeutic strategies and set realistic expectations for patients and families. Future research should involve extended longitudinal observations and a broad range of outcome measures, with a specific focus on identifying predictive and response biomarkers to optimise care across the entire SMA spectrum.

## Acknowledgments

We would like to acknowledge all the SMA patients and their families for participation in the CUIDAME registry; Carmen Baviera Ricart, Fernando Mora Tatay, Juan Carlos Leon Castro, Eugenia Ibañez Albert, Francisco Javier Sanz de Galdeano, Paula Lizandra, Silvia Albertos for evaluations and study coordination. We thank Marnee McKay, Amy Pasternak and Giorgia Coratti for instructions and source data for calculation of the FCR-composite score.

NM, RS, TS and JFVC are members of the European Reference Network for Rare Neuromuscular Diseases (ERN EURO-NMD).

## Declarations

### Authors’ contributions

KAG, DH and JFVC contributed to conception and design of the study. KAG and JFVC drafted the manuscript. DH performed statistical data analysis contributed to drafting a significant portion of the manuscript or figures. KAG, NCNG, NM, RS, TS, IPC, JFVC contributed to the clinical data acquisition and interpretation, critically revised and edited the manuscript.

JFVC had full access to all the data in the study and takes responsibility for the integrity of the data and the accuracy of the data analysis. All authors have approved the submitted version of the manuscript.

### Ethics approval

This study was approved by the Ethics Committee for Biomedical Research of La Fe Hospital (Valencia, Spain) 2019-027-1 and all participants provided informed consent.

### Availability of data and material

Anonymized data not published within this article will be made available by reasonable request from any qualified investigator

