## Supplementary figures and images for "Long-term adherence, safety and effectiveness of nusinersen in spinal muscular atrophy patients: a population-based study"

### Supplementary figure

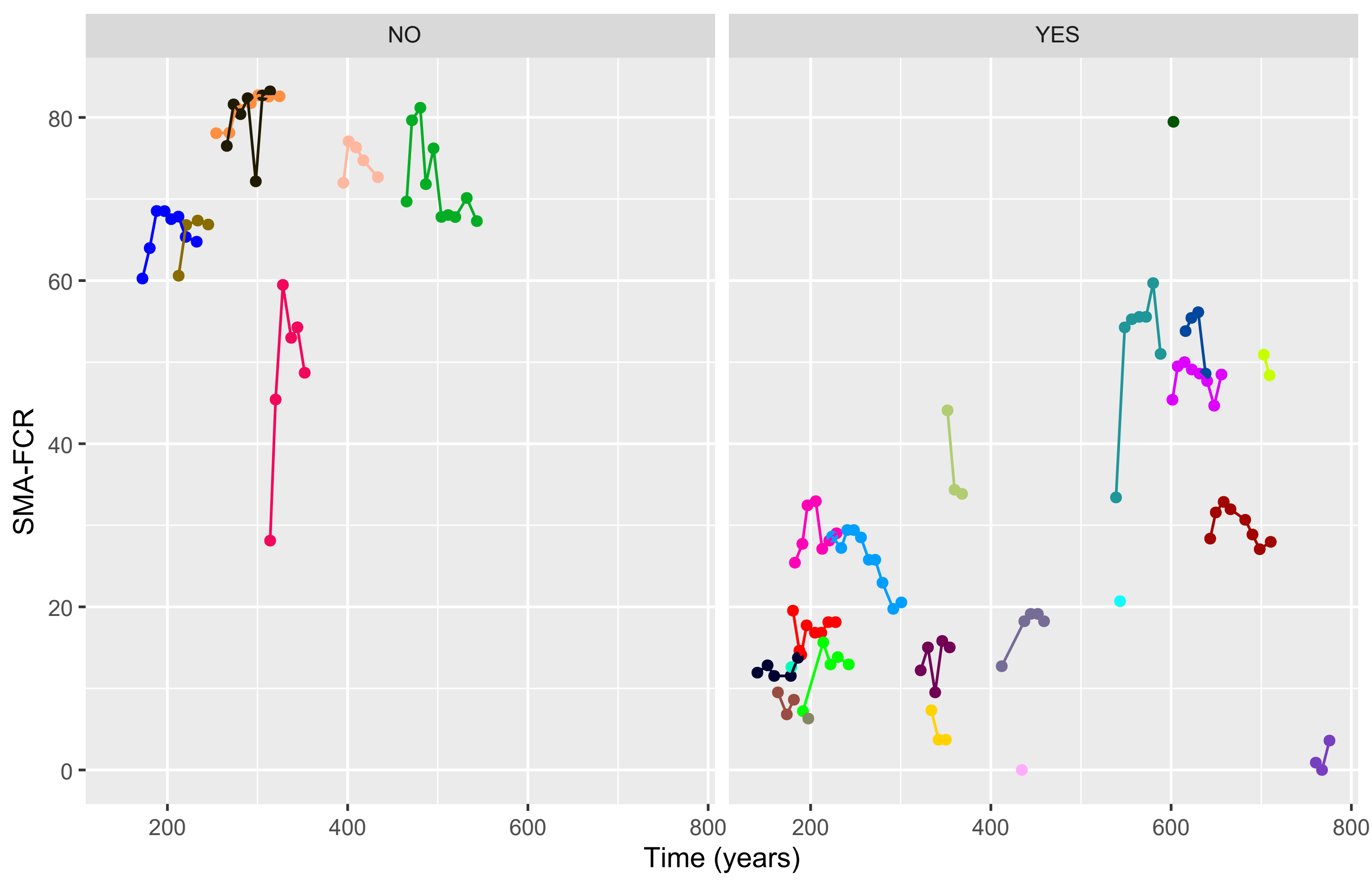
