## Supplementary file for "Long-term adherence, safety and effectiveness of nusinersen in spinal muscular atrophy patients: a population-based study"

Methodology

Spanish SMA treatment protocol criteria

In pediatric patients, inclusion criteria included all symptomatic patients and those presymptomatic with 0-3 copies. In adult patients, inclusion criteria included symptomatic patients with an objective impairment in the evaluation or scales and evidence of disease progression in the last 5 years (as per patient report or medical reports). Exclusion criteria were off-label use (including patients without lumbar access), patients with very severe impairment (minimal function and ≥16 hours of non-invasive ventilation -NIV-) and unfavourable risk-benefit ratio as per clinical judgement. In our region, patients not meeting criteria could also receive treatment, if clinically justified, on a case-by-case basis and after local approval.

Motor scales

CHOPINTEND, Hammersmith Functional Motor Scale - Expanded (HFMSE) and Revised Upper Limb Module (RULM) and 6-Minute Walk Test (6MWT) were used to evaluate patients at the appropriate age.

The HFMSE, which consists of 33 items, with a maximum of 66 points (higher scores indicating better function). This scale was originally designed for the assessment of high- functioning type 2 and 3 SMA patients, that is, sitters and walkers, although it shows floor effect in low-function sitters (Vázquez-Costa, Povedano, Nascimiento-Osorio, Escribano, García, et al., 2021). The RULM, which includes 20 items with a maximum score of 37 (higher scores indicating better function). Although it has been validated in both ambulant and non- ambulant patients, it shows a ceiling effect in up to a third of ambulant patients with SMA type 3 and a floor effect in a proportion of non- sitters (Vázquez-Costa, Povedano, Nascimiento-Osorio, Escribano, García, et al., 2021).

Results

| **Variables** | **exp.Estimate.** | **Lower.95.** | **Upper.95.** | **p** |
| --- | --- | --- | --- | --- |
| Intercept | 1.139 | 0.084 | 13.37 | 0.547 |
| Age | 1.028 | 1.011 | 1.055 | 1 |
| Male sex | 0.148 | 0.005 | 2.41 | 0.902 |
| SMA-FCR | 0.84 | 0.718 | 0.93 | 1 |

Supplementary Table 1. Bayesian model assessing the independent effect of age, sex and baseline SMA-FCR in the risk of nusinersen discontinuation in patients ≥12 years old.

| **Variables** | **Estimate** | **Std. Error** | **Lower 95** | **Upper 95** | **p** |
| --- | --- | --- | --- | --- | --- |
| Male sex | 2.007 | 8.416 | -14.866 | 18.818 | 0.593 |
| Time (months) | -0.017 | 0.02 | -0.056 | 0.02 | 0.815 |
| Maintained | -3.23 | 14.173 | -30.773 | 24.246 | 0.587 |
| Discontinuation | -7.908 | 13.885 | -34.877 | 19.882 | 0.72 |
| **Time:matained** | **0.098** | **0.032** | **0.036** | **0.162** | **1** |
| Time:discontinued | -0.013 | 0.027 | -0.067 | 0.042 | 0.682 |

Supplementary Table 2. Multivariable mixed beta model assessing the trajectories of SMA-FCR of the different subjects with time. In bold, statistically significant results.

Supplementary Figure 1

Individual SMA-FCR trajectories of patients sustaining and discontinuing nusinersen according to their age at baseline
